# Long-term performance assessment of fully automatic biomedical glottis segmentation at the point of care

**DOI:** 10.1101/2022.04.01.22273289

**Authors:** René Groh, Stephan Dürr, Anne Schützenberger, Marion Semmler, Andreas M. Kist

## Abstract

Deep Learning has a large impact on medical image analysis and lately has been adopted for clinical use at the point of care. However, there is only a small number of reports of long-term studies that show the performance of deep neural networks (DNNs) in such a clinical environment. In this study, we measured the long-term performance of a clinically optimized DNN for laryngeal glottis segmentation. We have collected the video footage for two years from an AI-powered laryngeal high-speed videoendoscopy imaging system and found that the footage image quality is stable across time. Next, we determined the DNN segmentation performance on lossy and lossless compressed data revealing that only 9% of recordings contain segmentation artefacts. We found that lossy and lossless compression are on par for glottis segmentation, however, lossless compression provides significantly superior image quality. Lastly, we employed continual learning strategies to continuously incorporate new data to the DNN to remove aforementioned segmentation artefacts. With modest manual intervention, we were able to largely alleviate these segmentation artefacts by up to 81%. We believe that our suggested deep learning-enhanced laryngeal imaging platform consistently provides clinically sound results, and together with our proposed continual learning scheme will have a long-lasting impact in the future of laryngeal imaging.

## Introduction

Laryngeal videoendoscopy is a major assessment tool to evaluate voice physiology qualitatively and quantitatively (Fig. 1). Especially for quantifying voice physiology, high-speed videoendoscopy (HSV) [1, 2] is an emerging technique that is able to visualize each glottal cycle with high spatial and temporal resolution. As the vocal folds, the main source of our voice, are vibrating hundreds of times each second, high-speed recordings with at least 4,000 frames per second (fps) are needed to accurately record this motion [1, 3].

**Fig 1.**
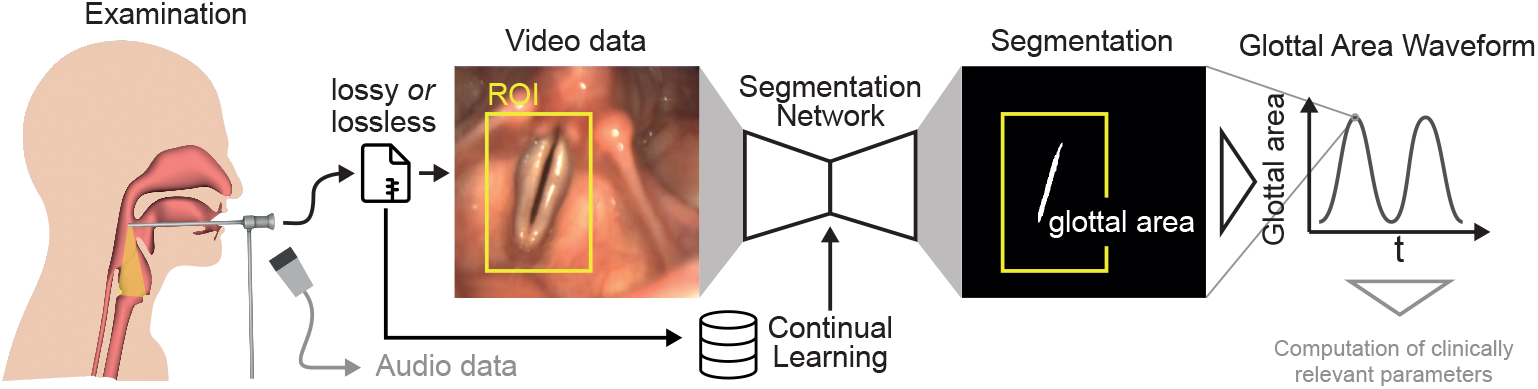
Data acquisition and analysis workflow. Each examination yields video and audio data, where the video data is stored either lossy or lossless compressed. In this study, we evaluate which data source and if cropping the video data to an ROI is sufficient for reliable glottis segmentation using previously proposed clinically optimized segmentation networks. The glottal area is computed for each video frame and plotted across time yielding the glottal area waveform (GAW), which is crucial for computing clinical relevant parameters. We investigate in this study how collected data can be used to allow constant fine-tuning of the segmentation network using continual learning schemes.

The glottal area, the opening between the two vocal folds, is a good proxy for assessing the oscillation behavior [1, 4]. Therefore, many works have focused on the segmentation of the glottal area (Fig. 1), especially avoiding any manual intervention [4, 5]. This critical step is one of the main bottlenecks of the data analysis pipeline and has been the reason why HSV is barely applied in the clinic, because fully automatic data analysis solutions have not been around [1]. Lately, it has been shown that deep neural networks (DNNs) are highly suitable for solving this task [6–9]. These glottis segmentation DNNs could be further optimized towards clinical applicability with barely sacrificing segmentation accuracy [6]. Together with the recent development of an open source HSV system, namely OpenHSV, latest hardware and software components were introduced to the clinic [10] that features these optimized DNNs for clinical use. However, there is no record how these DNNs perform actually in a clinical environment, as they have been validated on limited and selected data.

In this work, we report the performance of the AI-powered OpenHSV system together with the DNN in a two-year clinical environment. Our main contributions are summarized as follows: (1) we describe for the first time the overall image quality distribution during a two-year period of clinical use, and if lossy data compression is suitable for data storage and subsequent data analysis, (2) give an unbiased performance evaluation of previously proposed optimized DNNs for clinical use for different data origins and (3) employ a continual learning and fine-tuning strategy to allow continuous integration of novel data into the DNN. Taken together, we strongly believe that our study provides trust and shows reliability for the OpenHSV system positively impacting future clinical adaptation.

## Methods

### OpenHSV system

We are using the OpenHSV platform introduced in [10]. Patients are routinely examined using a rigid endoscope equipped with a high-speed camera (IDT CCmini-1540) running at 4,000 fps. Illumination is granted by a high-power LED light source (Storz 300 W LED). Each recording is at least 1,000 frames long and contains synchronously acquired video and audio data. We further record patient metadata consisting of the patient’s age, gender, and condition. For each video, we save two files encoded with the h.264 codec: (1) lossy compression using common settings for video and (2) lossless compressed data to recover the original recorded raw data. For lossy compression, we use the libx264 codec, the yuv420p pixel format and set the quality to 5 resulting in a varying bitrate. For lossless compression, we used the libx264rgb codec, the rgb24 pixel format, set the ‘-crf’ flag to 0 and used the ultrafast preset.

### Patients

We assessed a total of 583 recordings acquired between November 2019 and November 2021. All recordings are done routinely in the clinic and are performed according to local regulations (Ethikkommission FAU Erlangen-Nürnberg, #290 15). We first selected only those recordings that featured a complete set of data, such that 295 recordings remained. Next, we manually investigated the data quality. We ranked each video on an ordinary scale: 0 (insufficient), 1 (okay), 2 (excellent). Videos ranked 0 were showing insufficient data quality, such as non-visible glottis or foggy videos, and were discarded. Finally, 267 recordings from 202 unique patients remained and were subjected to further analyses (Fig. 2A). We report the frequency of recordings across the last two years in Fig. 2B. Additionally, COVID-19 cases for Germany were provided as a reference how data generation was affected by lock-downs. The number of recordings were higher before the first lock-down, but their fluctuation remained constant when the general clinical activity was restored. The age distribution of the patients is largely spread (Fig 2C), where the mean age is 47.2 *±* 20.2 (std) years. The reported gender for the analyzed subjects is 31.2% male, 66.8% female and 2.0% had no further specified gender. The average file size for lossless and lossy recordings was 2.06 *±* 1.27 GB and 8.97 *±* 5.16 MB, respectively (see Fig. 2D). As the lossy files have an around 235-times lower file size than the lossless compressed counterparts, we investigated in this study if the lossy compression has an impact on the segmentation performance.

**Fig 2.**
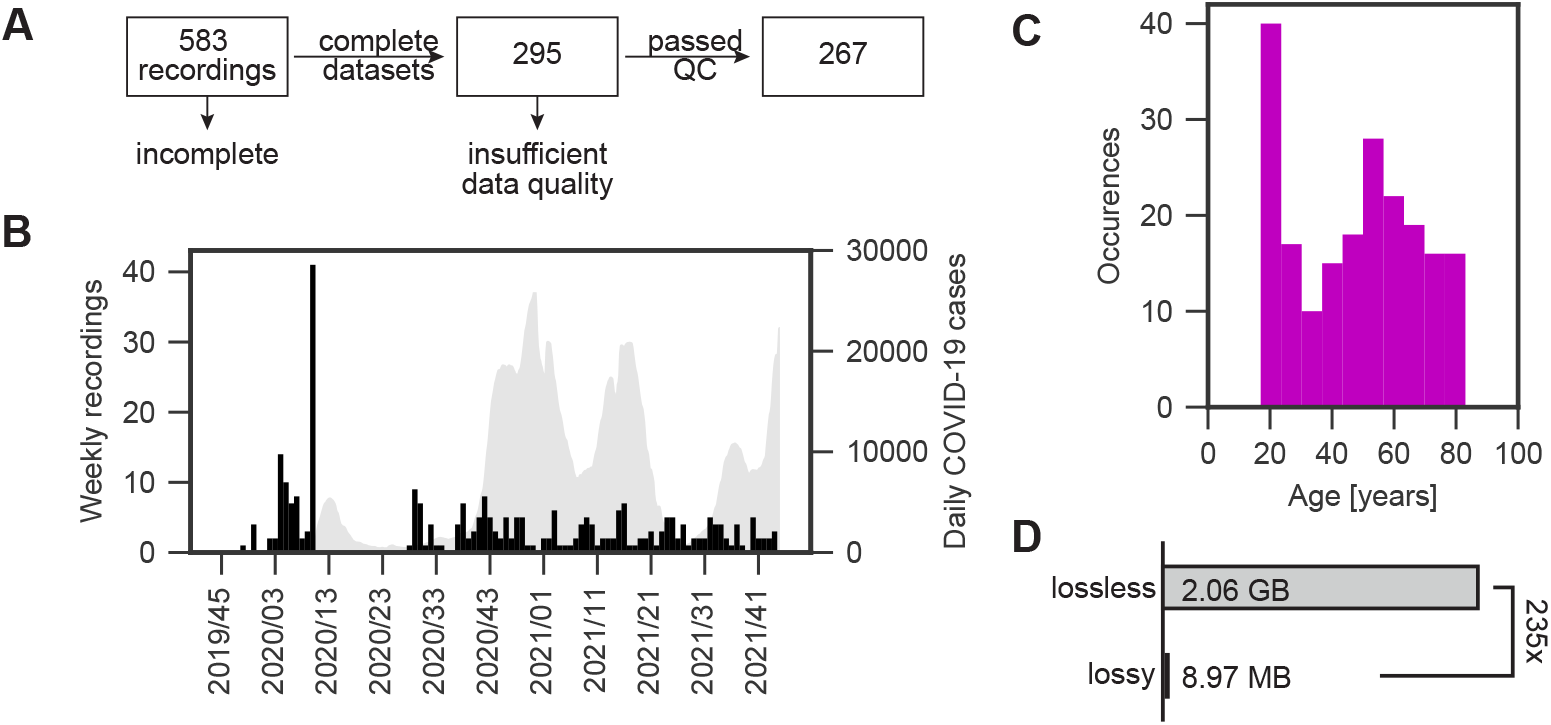
Acquired OpenHSV data overview. A: From initial 583 recordings, we excluded datasets that are missing any kind of data. From these, we excluded ones with insufficient data quality, such as occluded glottis. B: Frequency of recordings across the two year time frame is shown in black. The Germany COVID-19 cases are shown as a 7-day-average in gray. C: Age distribution across selected recordings from A. D: Comparison of file size between lossy and lossless compression.

### Image quality assessment

As we saved the data in two different compression modes, namely lossy and lossless, we evaluated if there are any compression artefacts. First, we investigated the dynamic range of the image, where 0 is no dynamic range and 1 is full dynamic range by computing the normalized absolute difference of the minimum and maximum pixel value. We determined the no-reference, completely blind image quality score “Natural Image Quality Evaluator” (NIQE) [11] to provide an unbiased score for objective image quality assessment. We further assessed full reference metrics, such as peak signal to noise ratio (PSNR) and structural similarity index metric (SSIM) [12] to compare lossy compression against the lossless stored data. We use respective implementations of the metrics in scikit-video (NIQE) and scikit-image (PSNR, SSIM).

### Deep Neural Network

The OpenHSV system is shipped with a deep neural network (DNN) based on the U-Net architecture [13], that was optimized for clinical use as described previously [6]. The used DNN is openly available on the OpenHSV Github account at https://github.com/anki-xyz/openhsv. The training process is described in [10]. Briefly, the DNN is setup in TensorFlow 2.2 and the high-level Keras package. The DNN was pretrained in a supervised fashion on the full training dataset (55,750 images) of the open benchmark for automatic glottis segmentation (BAGLS, [7]). The pretrained network has never been exposed to OpenHSV data during training.

### Region of interest

A rectangular region of interest (ROI) was drawn manually for each recording. We saved the ROI coordinates for further use in JSON format. Each ROI was adjusted as such that the width and height is divisible by 32 to ensure proper DNN propagation.

### Segmentation

Lossy or lossless compressed endoscopic frames were first converted to grayscale by extracting the luminance channel using standard procedures, as it has been shown that color information is not essential for glottis segmentation [7]. In some experiments, only an ROI around the vocal folds was used for inference. The input image intensity was normalized between -1 and 1. The segmentation mask gained from the DNN provides values in the range of 0 (background) and 1 (glottis) by a sigmoid function in the output layer. For further use and due to memory limitations, we multiplied the predicted segmentation masks by 255 in order to save the data in uint8 data format. The glottal area waveform (GAW) is computed by summing the segmented pixels in each frame for every timepoint of a given recording (see Fig. 1).

### Continual training

We performed retrospective continual training on the original OpenHSV segmentation DNN. We preprocessed new images as described above and used two continual learning strategies: We either selected a fixed time period for data collection (7, 14, 30 days) or a fixed video quantity (every 10, 20 or 40 videos, see also Fig. 5A). At each continual learning point, we trained the model for ten epochs using the previously predicted segmentation masks as ground truth for the training process. We chose a low learning rate of 10^*−*6^ combined with the Adam optimizer to fine-tune the model. After each continual training step, we evaluated the occurrence of artefacts by visual assessment (number of artefacts, shown in Fig. 5B and S3 Fig) and computed the achieved Intersection over Union (IoU, [14]) score (Fig. 5C,D). The IoU score is a measure how well the prediction segmentation mask overlaps with the ground-truth segmentation mask and ranges between 0 (no overlap) and 1 (perfect overlap).

## Results

### Recording quality is consistent across time

To evaluate the performance of the segmentation DNN, we first assessed the overall image quality for both, lossless and lossy recordings, as this is a major confounding source for segmentation success. First, we determined the maximum dynamic range of each given image. We found that the maximum dynamic range is constantly high for both, lossless and lossy recordings (Fig. 3A, left panel), however, the dynamic range is significantly lower for lossy recordings as for lossless recordings (paired Student’s t-test, p <0.01, Fig. 3A, right panel). Using a complete blind, non-reference metric, the NIQE score [11], we could show that lossless recordings have lower, therefore better NIQE scores until beginning of 2021 (Fig. 3B), afterwards the NIQE scores were highly overlapping. In [10], a mean NIQE for the OpenHSV system of 13.19 was reported, showing that image quality has been consistent since the clinical introduction of the system.

**Fig 3.**
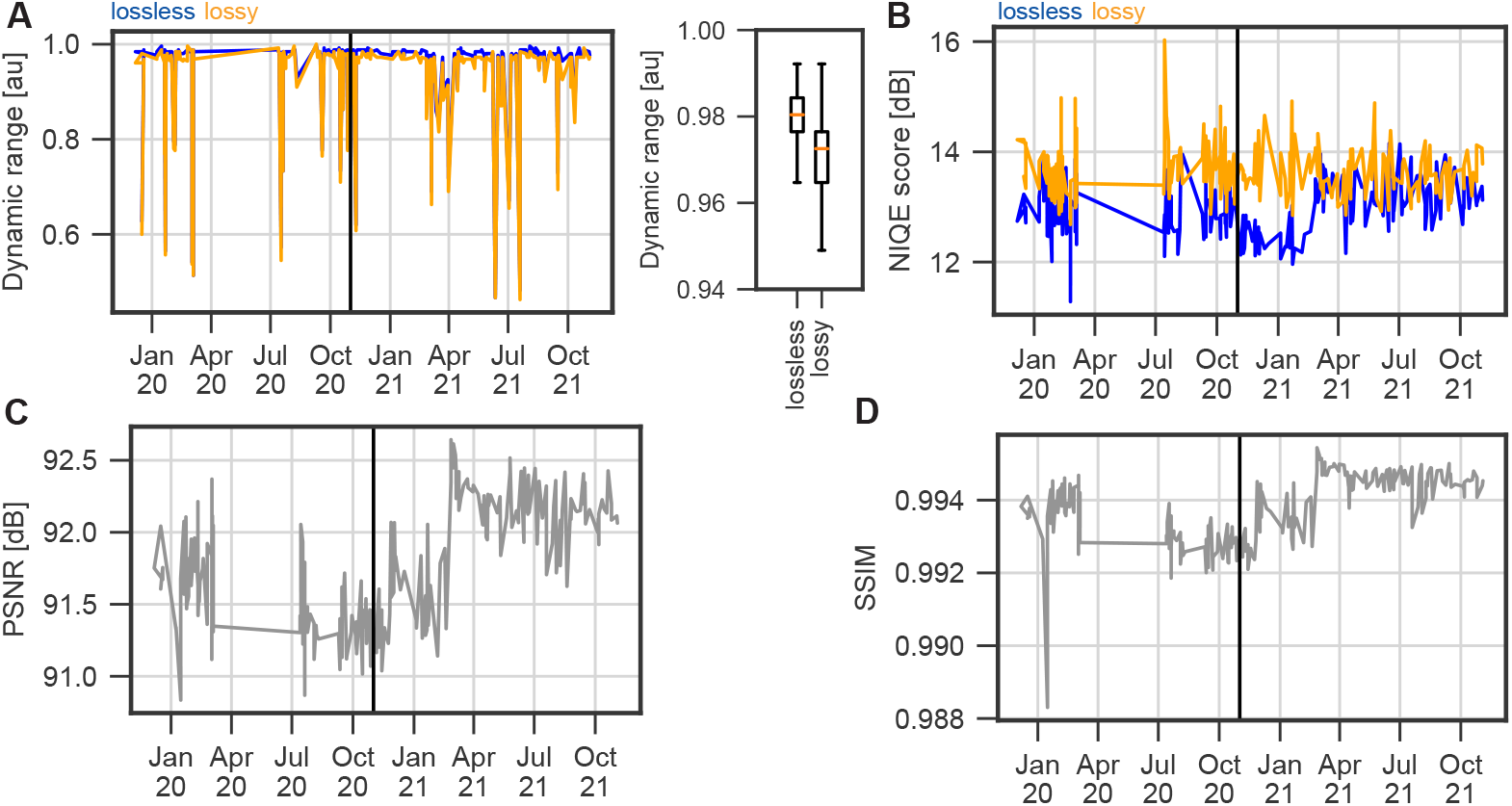
Image quality is relatively stable across time. A: Dynamic range per recording. Lossless compressed data in blue, lossy in orange. Boxplots show the 95-percentile of the data. B: NIQE score for lossy (orange) and lossless (blue) compressed videos across time. C: Peak-Signal-To-Noise-Ratio (PSNR) across time. D: Structural Similarity Index Measure (SSIM) across time.

To compare images retrieved from lossy recordings to their lossless counterparts, we relied on two reference metrics, peak-signal-to-noise-ratio (PSNR) and structural similarity index measure (SSIM). PSNR has a constant high value above 90 dB, suggesting a high-quality compression (Fig. 3C). In contrast to PSNR, SSIM also takes the perceptual change in structural information into account. In Fig. 3D we show that the lossy compressed videos nevertheless are in almost perfect agreement with the lossless reference.

In summary, we found that the image quality has single outliers, but overall is highly consistent across time. Further, high PSNR and SSIM values indicate an overall accurate conversion from lossless to lossy image content and high quality in lossy compressed video.

### Segmentation performance is not affected by lossy data compression

To further evaluate the performance of the segmentation DNN, we manually annotated the glottal area in the first 100 frames of 20 randomly selected videos, half of which were rated as quality 1 (okay) and the other half as quality 2 (excellent). Using this ground truth and the predicted segmentation masks by the DNN, we computed the Intersection over Union (IoU) for each frame across all videos. We found that lossy and lossless saved videos achieve a similar performance with a median IoU of 0.756/0.742 and 0.770/0.768 for segmentation masks computed with and without ROI, respectively (Fig. 4B). These IoU values are comparable to other works [6], where IoU scores between 0.741 and 0.769 were achieved, and are sufficient for clinical reliability.The use of an ROI enhances the segmentation speed as smaller images are used, however, this leads to worse results. We hypothesize that this is caused due to the loss of global spatial information. We further mined the computed IoU scores to determine why very low IoU scores are obtained. Fig. 4C shows that the low IoU scores emerge with a small segmented area, i.e. when the glottis is closed. This is in line with previous reports [7], and has a negligible effect on the data analysis. We next investigated if any configuration has an impact on the clinically relevant glottal area waveform (GAW) signal. We were able to confirm that all combinations despite their deviation in the IoU score have little to no affect on the GAW, as they do not deviate (S1 Fig A), and almost perfectly correlate (S1 Fig B), important for downstream computation of quantitative parameters.

**Fig 4.**
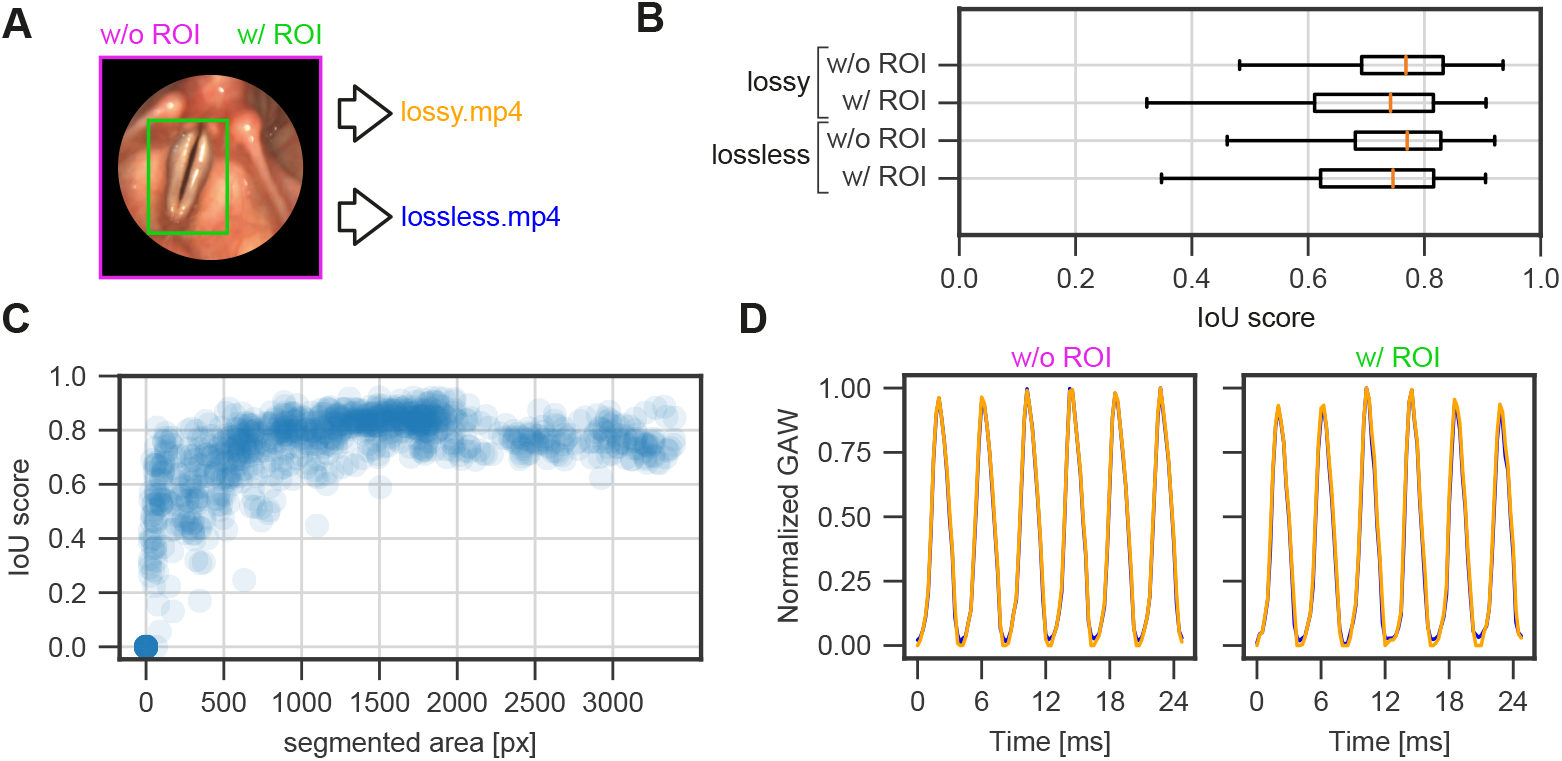
Lossy compression does not affect segmentation performance. A: Input data for DNN inference. Either full frame (magenta) or ROI-constrained (green) image data were used for inference. Image data were stored either lossless or lossy compressed as .mp4-files (see Methods). B: Intersection over Union (IoU) score per configuration shown in A. C: Dependency of IoU scores on the segmented glottal area. D: Exemplary glottal area waveform for full frame (w/o ROI) and ROI-based (w/ ROI) segmentations. Lossy and lossless compressed data are plotted in orange and blue, respectively.

### Continual training for DNN fine-tuning improves performance

Despite the fact that we gained mostly successful and accurate segmentations, we found for a minority of videos (9%) two common issues: artefacts in the segmentation masks and empty segmentation masks (S2 FigA). We hypothesized that continuous integration of new, system-specific data using continual training [15, 16] is increasing the DNN performance. Here, we evaluated two strategies for integrating new data (Fig. 5A). We either used a fixed time period or fixed quantities of videos. We used artefact-free, full-frame, lossy compressed videos for continual training, as full frames resulted in higher IoU scores (Fig. 4A). We found that all strategies were able to reduce the number of artefacts after only the first iteration of continual training and removed artefacts on average by 38-48% (Fig. 5B). Additionally, the more data is used for each training step, the higher the impact on artefact removal, and that a fixed quantity is preferable to a fixed time period (Fig 5B). In particular, when determining the median, i.e. what is the reduction in 50% of all cases, we found that the fixed amount of 40 videos has the best overall performance of reducing the artefacts of by 48%. In general, we can confirm that a fixed data amount seems to be preferable to a fixed time frame, as the latter also shows an unstable performance behavior across continual learning epochs (S3 Fig). To further quantify this effect more objectively, we annotated the first 30 frames of each artefact video to compute IoU scores after each continual learning step (Fig 5C-D). Each strategy was able to increase the IoU score already after the first continual learning iteration, whereas a fixed data amount has a more stable and constant performance (Fig 5C) compared to a fixed time interval (Fig 5D), similar to our observations with manually scored artefacts (S3 Fig). Looking at the strategy of using fixed time periods, it is clear that the IoUs do not change significantly during the first COVID-19 wave, which again shows that the use of fixed quantities is preferable to fixed time periods. Taken together, our data shows that the segmentation DNN is not only able to quickly adapt to new data, but also that continual learning is an important feature in using DNNs in a clinical context.

**Fig 5.**
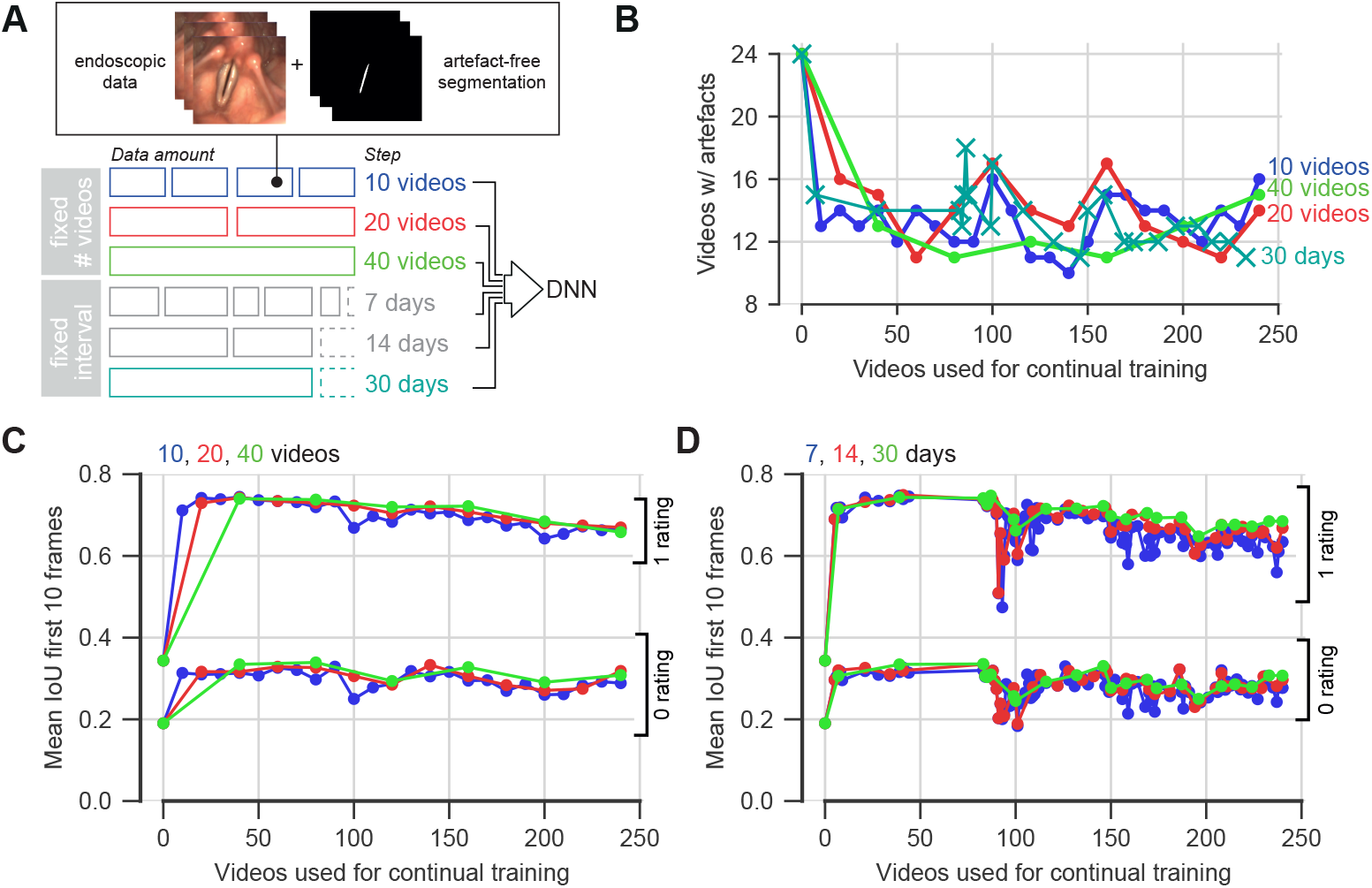
Continual training alleviates segmentation artefacts. A: Continual training strategies. B: Reduction of artefacts depending on continual learning strategy. C: IoU score of the first ten frames of the evaluated data after continual learning with a fixed amount of videos. D: Same as in C, but with a fixed time interval.

## Discussion

Laryngeal high-speed videoendoscopy is a major tool in quantifying laryngeal physiology. In this study, we show that clinically optimized DNNs have an overall high performance due to a constantly high data quality and a well pre-trained, generalized DNN. Although the DNN has never be trained on this system’s data, we still gain in most (91%) cases satisfactory results. We further show that lossy compressed videos are on par with lossless compressed videos in terms of segmentation performance (Fig. 4). Using continuous integration of novel data, we can also show that the DNN is able to adapt such that previously identified artefacts are reduced and ensure a constantly improving segmentation environment (Fig. 5).

### Acquisition circumstances as potential confounders

We could show that the analyzed data has a relatively high image quality throughout the analyzed time frame (Fig. 3). However, other systems have been shown to suffer from low lighting, which can be rescued with Deep Learning methods [17]. As the data acquisition is a manual procedure, motion of the patient or the examiner can be a confounder. Motion correction techniques were proposed [18, 19] that can be used as a pre-processing step that have not been employed in this analysis.

### Comparison to other glottis segmentation platforms

In this study, we investigated the performance of a single DNN. Offline image analysis platforms, such as the Glottis Analysis Tools [19] (GAT), can further serve as a reference for segmentation performance and may allow segmentations of higher quality. In comparison, the analyzed OpenHSV DNN has similar performance as the smallest GAT neural network, however, larger and more elaborate networks have superior performance (S1 Table). Notably, this effect is largely compensated by our continual training scheme (Fig. 5). Nevertheless, the average IoUs obtained in this study are in an acceptable range (0.742-0.770) that do not impact the clinical soundness of downstream quantitative parameter computation as shown previously [6, 10].

### Continual learning strategies

Our proposed continuous integration of more data is straightforward and effortless. We have seen that using a fixed data mount is beneficial to a fixed time interval, which is maybe due to the irregular patient stream (Fig. 2). We further observe a decline in IoU scores across data and time (Fig. 5C-D) and a tendency to more artefacts after integrating a large amount of data (Fig. 5B, S3 Fig). To counteract catastrophic forgetting [15], a common problem in continual learning schemes, additional precautions can be introduced, such as integrating the BAGLS dataset [7] in the continual learning training dataset or testing on previous artefact-free images if the current model performs better than before. Combining both aforementioned examples would incorporate external and internal regularization factors that are maybe beneficial in a more elaborate continual learning paradigm. In addition, we have not investigated how more sophisticated human-in-the-loop strategies [20], such as manual segmentation and retraining, perform. With a wider adoption of OpenHSV, we also believe that federated learning techniques [21] will further boost the DNN segmentation performance.

## Data Availability

Primary data cannot be shared publicly because of patient data protection and privacy measurements. Data are available upon reasonable request from the corresponding author in close exchange with the ethics committee of the University Hospital Erlangen for researchers who meet the criteria for access to confidential data. However, we do share the code that has been used to analyze the data and the deep neural network such that our results can be confirmed independently.

## Supporting information

**S1 Fig. GAWs are highly correlated across compression methods**. A: Distribution of mean absolute error (MAE) across compression modes segmented w/ ROI (green) or w/o ROI (magenta). B: Distribution of the Pearson’s correlation coefficient across compression modes segmented w/ ROI (green) or w/o ROI (magenta).

**S2 Fig. Continual learning has an effect on erroneous segmentations (upper panels) and on failed segmentations (lower panels)**.

**S3 Fig. Artefacts decay with continual learning**. A. Videos with artefacts according to rating (0=large artefacts, failed segmentations, 1=small artefacts, erroneous segmentations) for continual learning with a fixed amount of videos, namely 10 (yellow), 20 (green) or 40 (magenta). B. Same as panel A, but with fixed time interval of 7 (yellow), 14 (green) or 30 (magenta) days.

**S1 Table. Larger pre-trained DNNs perform slightly better in glottal segmentation before fine-tuning our model**. Number of Artefacts and Intersection over Union (IoU) for several deep neural networks.

## Acknowledgments

This work was supported by the German Research Council (DFG) under grant no. SCHU 3441/3-2.

## Notes

### Competing Interest Statement

The authors have declared no competing interest.

### Funding Statement

This work was funded in part by the German Research Foundation (DFG, https://www.dfg.de/) under grant no. SCHU 3441/3-2 (to AS).

### Author Declarations

The ethics committee of the Friedrich-Alexander-University and the University Hospital Erlangen approved the study (approval number #290_15).

